# SARS-CoV-2 infection induces cross-reactive autoantibodies against angiotensin II

**DOI:** 10.1101/2021.11.02.21265789

**Authors:** Priscilla S. Briquez, Sherin J. Rouhani, Jovian Yu, Athalia R. Pyzer, Jonathan Trujillo, Haley L. Dugan, Christopher T. Stamper, Siriruk Changrob, Anne I. Sperling, Patrick C. Wilson, Thomas F. Gajewski, Jeffrey A. Hubbell, Melody A. Swartz

## Abstract

Patients infected with the severe acute respiratory syndrome coronavirus-2 (SARS-CoV-2) can experience life-threatening respiratory distress, blood pressure dysregulation and thrombosis. This is thought to be associated with an impaired activity of angiotensin-converting enzyme-2 (ACE-2), which is the main entry receptor of SARS-CoV-2 and which also tightly regulates blood pressure by converting the vasoconstrictive peptide angiotensin II (AngII) to a vasopressor peptide. Here, we show that a significant proportion of hospitalized COVID-19 patients developed autoantibodies against AngII, whose presence correlates with lower blood oxygenation, blood pressure dysregulation, and overall higher disease severity. Anti-AngII antibodies can develop upon specific immune reaction to the SARS-CoV-2 proteins Spike or RBD, to which they can cross-bind, suggesting some epitope mimicry between AngII and Spike/RBD. These results provide important insights on how an immune reaction against SARS-CoV-2 can impair blood pressure regulation.

## INTRODUCTION

Severe acute respiratory syndrome coronavirus-2 (SARS-CoV-2), the causative virus of coronavirus disease 2019 (COVID-19), infects cells by binding to angiotensin-converting enzyme-2 (ACE-2) via the receptor-binding domain (RBD) of its Spike protein. ACE-2 is an enzyme expressed on the surfaces of alveolar epithelial cells and vascular endothelial cells (*1*), among others, that plays an important role in regulating blood pressure by converting the vasoconstrictive peptide angiotensin II (AngII) to the vasopressor peptide angiotensin-(1-7) (*2, 3*). It is known that SARS-CoV-2 infection can lead to dysregulation of vascular tension, endothelial inflammation and enhanced thrombosis, presumably through enhancing endocytosis of ACE-2 and thereby lowering its cell-surface presence (*4, 5*).

Here, we sought to explore whether SARS-CoV-2 infection might induce auto-antibodies against the peptide AngII. We hypothesized that the simultaneous binding of SARS-CoV-2 and AngII to ACE-2 might lead to their co-phagocytosis by antigen-presenting cells, thus providing a strong immune adjuvant (the virus molecules) to the self-peptide AngII, leading to an anti-AngII autoimmune response (*6, 7*). Moreover, we asked whether some epitope mimicry might exist between a domain on the Spike protein and AngII, based on their shared binding to ACE-2. Importantly, the induction of anti-AngII antibodies in COVID-19 patients, if it occurs, could interfere with AngII processing by ACE2 and signaling to its receptors, potentially contributing to the dysregulation of vascular tension and worsening acute respiratory distress syndrome(*8, 9*).

We conducted observational studies using serum of hospitalized COVID-19 patients and determined that such autoantibodies are indeed induced, independently of anti-RBD levels. Instead, their presence and levels were strongly correlated with blood pressure dysregulation and poor oxygenation. We finally demonstrated cross-reactivity of some antibodies between AngII and the Spike protein, suggesting immune epitope homology between these molecules.

## RESULTS AND INTERPRETATION

We began by assessing the presence of IgG antibodies against AngII in the plasma of 221 subjects, among which 115 were hospitalized COVID-19 patients convalescent from a SARS-CoV-2 infection, 58 were control donors (i.e. non-SARS-CoV-2 infected and non-hypertensive), and 48 were hypertensive non-SARS-CoV-2 infected donors. Surprisingly, we found that a substantial proportion, 63% (73/115), of the COVID-19 patients had positive levels of anti-AngII autoantibodies, as determined by an ELISA absorbance greater than 3 standard deviations above the mean of the control donor group **(Fig. 1A)**. Of these, 53% (39/73) had high levels, defined as greater than twice the positive threshold. In contrast, only one control donor (1.7%) was anti-AngII positive, with a level just above the positive limit. Patient age, sex, and body mass index (BMI) were not significantly correlated with anti-AngII positivity **(Fig S1A-D)**, although older patients trended towards increased levels of anti-AngII (p=0.064 by Spearman correlation, **Fig. S1A**). Additional trends suggested that female COVID-19 patients were more likely to develop high anti-AngII levels (42% vs. 25% of males, **Fig. S1B**) as well as patients with BMI ≥ 25 (38% vs. 20% of patients with normal BMI, **Fig. S1C, D**).

**Figure 1.**
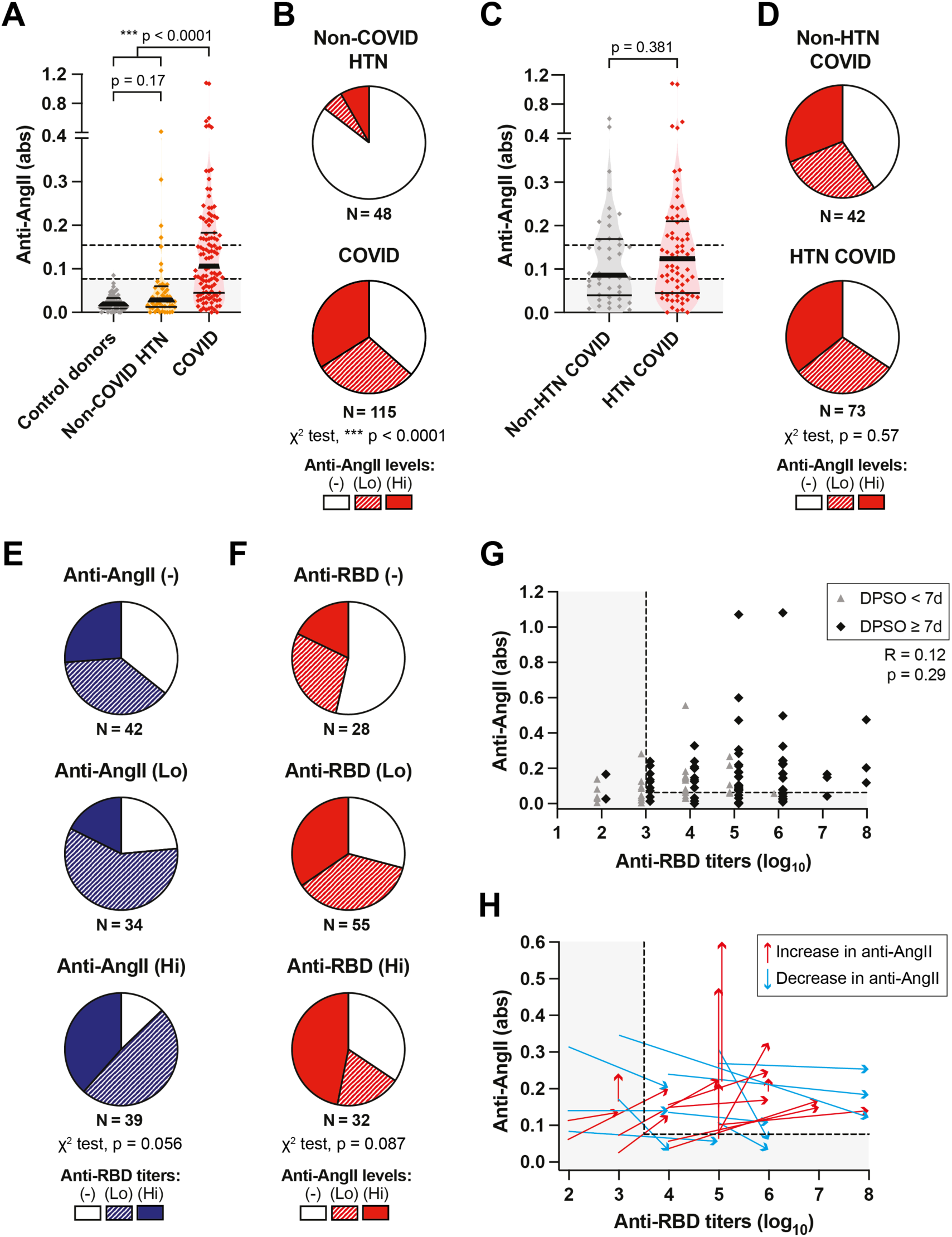
SARS-CoV-2 infection induces anti-AngII autoantibodies. The plasma of SARS-CoV-2 convalescent hospitalized patients (COVID, N=115) was analyzed for the presence of anti-AngII and anti-RBD by ELISA. Among them, 63% had with pre-existing hypertension (HTN COVID, N=73). Results were compared to non-COVID hypertensive donors (Non-COVID HTN, N=48) or control donors (N=58). Anti-AngII levels are displayed as the signal absorbance measured in plasma diluted at 1:100. Anti-RBD titers are displayed as log_10_ values. (Hi = high; Lo = low; (-) = negative; gray thresholds = limits for anti-AngII or anti-RBD positivity) **(A)** Levels of anti-AngII antibodies in the plasma of COVID patients as compared to non-COVID HTN donors and control donors (median ± IQR, Kruskal-Wallis test with Dunn’s post-test). An absorbance ≥ 0.077 and ≥ 0.154 indicates the limit for anti-AngII positivity and high levels, respectively. **(B)** Proportion of non-COVID HTN and COVID donors that have high (Hi), low (Lo) or negative (-) levels of anti-AngII (χ^2^ test). **(C)** Levels of anti-AngII in COVID patients with or without pre-existing HTN (Mann-Whitney test). **(D)** Proportion of non-HTN COVID and HTN COVID patients that have high (Hi), low (Lo) or negative (-) levels of anti-AngII (χ^2^ test). **(E)** Proportion of patients with high, low or no titers of anti-RBD among patients having high, low or negative levels of anti-AngII (χ^2^ test). **(F)** Proportion of patients with high, low or negative levels of anti-AngII among patients having high, low or no titers of anti-RBD (χ^2^ test). **(G)** Correlation between anti-AngII levels and anti-RBD titers in COVID patients. Patients for which the plasma was analyzed at or after 7 days post-symptoms (DPSO) are indicated in black, whereas those analyzed before 7 DPSO are in grey (Spearman correlation on datapoints ≥ 7 DPSO). **(H)** Co-variations of anti-AngII and anti-RBD levels between 1-10 DPSO (tail of the arrow) and 11-20 DPSO (tip of the arrow) matched per patient (arrow, N=25).

Since a majority of the hospitalized COVID-19 patients in this cohort had pre-existing hypertension (COVID HTN, 64%), we measured anti-AngII levels in plasma from hypertensive donors taken prior to the pandemic (non-COVID HTN) to determine whether autoantibodies against AngII could have been pre-existing in HTN patients. Interestingly, we detected anti-AngII antibodies in 15% of these hypertensive donors **(Fig. 1A, B)**, which was far less than in COVID-19 patients. Furthermore, when comparing the HTN vs. non-HTN COVID-19 patients, the levels of anti-AngII were similar **(Fig. 1C)** as well as the percentages with positive levels (65% and 60%, respectively; **Fig. 1D**). These results indicate that infection with SARS-CoV-2 promotes the development of anti-AngII IgG antibodies in a majority of patients that developed severe disease (i.e., required hospitalization), regardless of whether they had pre-existing hypertension.

Next, we asked whether the presence and levels of anti-AngII antibodies correlated with those of antibodies directed against the receptor-binding domain (RBD) of the virus’ Spike protein. As expected, most patients (76%) developed positive levels of anti-RBD antibodies, here considered above a total IgG titer of 3 based on healthy control levels **(Fig. S2A)**. However, correlations between anti-RBD levels and anti-AngII levels were only modest; in fact, a majority of anti-AngII(-) patients developed antibodies against RBD **(Fig. 1E)**, and patients with high anti-RBD titers did not necessarily develop anti-AngII antibodies (34% remained anti-AngII(-)) **(Fig. 1F)**. In addition, although patients positive for one antibody were more likely to be positive for the other, this effect was not statistically significant (p=0.056 and p=0.087 using χ^2^ tests comparing the proportion of anti-AngII(+) patients across the anti-RBD (-/Lo/Hi) groups, and vice-versa). Similar results were observed when considering the average antibody levels instead of proportions of positive patients. Despite a slight increase in anti-AngII levels along with anti-RBD levels **(Fig**.**S2B, S2C)**, no significant correlation was observed between them when analyzing the plasma of patients collected after 7 days post-symptom onset (DPSO), which is the approximate time needed to observe a new IgG response in blood upon primary infection (**Fig. 1G**, p=0.29 by Spearman test).

We then questioned when anti-AngII antibodies developed relative to anti-RBD antibodies after SARS-CoV-2 infection by comparing their levels between early vs. later times (1-10 vs. 11-20 DPSO) in the 25 anti-AngII(+) patients for which these timepoints were available. From early to late, some patients showed increases in both anti-RBD and anti-AngII (**Fig. 1H** in red, 15/25), while others showed increases in anti-RBD accompanied by decreases in anti-AngII (**Fig. 1H** in blue, 10/25). Between these times, the increase in anti-RBD levels reached statistical significance **(Fig. S2D)**, as expected, whereas the increase in anti-AngII levels did not (**Fig. S2E**, p=0.18 by Wilcoxon matched pairs sign rank test). Moreover, anti-AngII antibodies were still detected in 11 patients at >45 DPSO (**Fig. S2F**), suggesting that either anti-AngII antibodies are long-lasting, or that they are maintained over a long-lasting symptomatic period in these patients. Together, these data suggest that antibodies levels generated against AngII are not necessarily correlated with those against RBD.

We next studied the development of anti-AngII IgG in mice upon exposure to SARS-CoV-2 antigens (note that murine and human AngII are 100% homologous). We vaccinated mice at 0 and 21 days with 10 μg Spike or RBD, admixed with either (i) the Toll-like receptor (TLR) 4 agonist monophosphoryl lipid A in alum (MPLA/Alum, mimicking the AS04 clinical adjuvant), (ii) AddaS03 (a formulation of composed of α-tocopherol, squalene and polysorbate 80 in an oil-in-water nano-emulsion, mimicking the AS03 clinical adjuvant), (iii) the TLR9 agonist CpG-B, or (iv) no adjuvant. None of the naïve mice (0/25) had detectable anti-AngII IgG **(Fig. 2A)**. In contrast, 24% (6/25) of the mice vaccinated with RBD + MPLA/Alum and 27% (4/15) of mice vaccinated with Spike + MPLA/Alum developed antibodies against AngII. Interestingly, none of the mice vaccinated with Spike + AddaS03 (0/9) or with RBD + CpG (0/10) developed anti-AngII antibodies, as well as none of the mice vaccinated with unadjuvanted RBD or Spike (0/10 each), or with MPLA/Alum lacking antigen (0/20).

**Figure 2.**
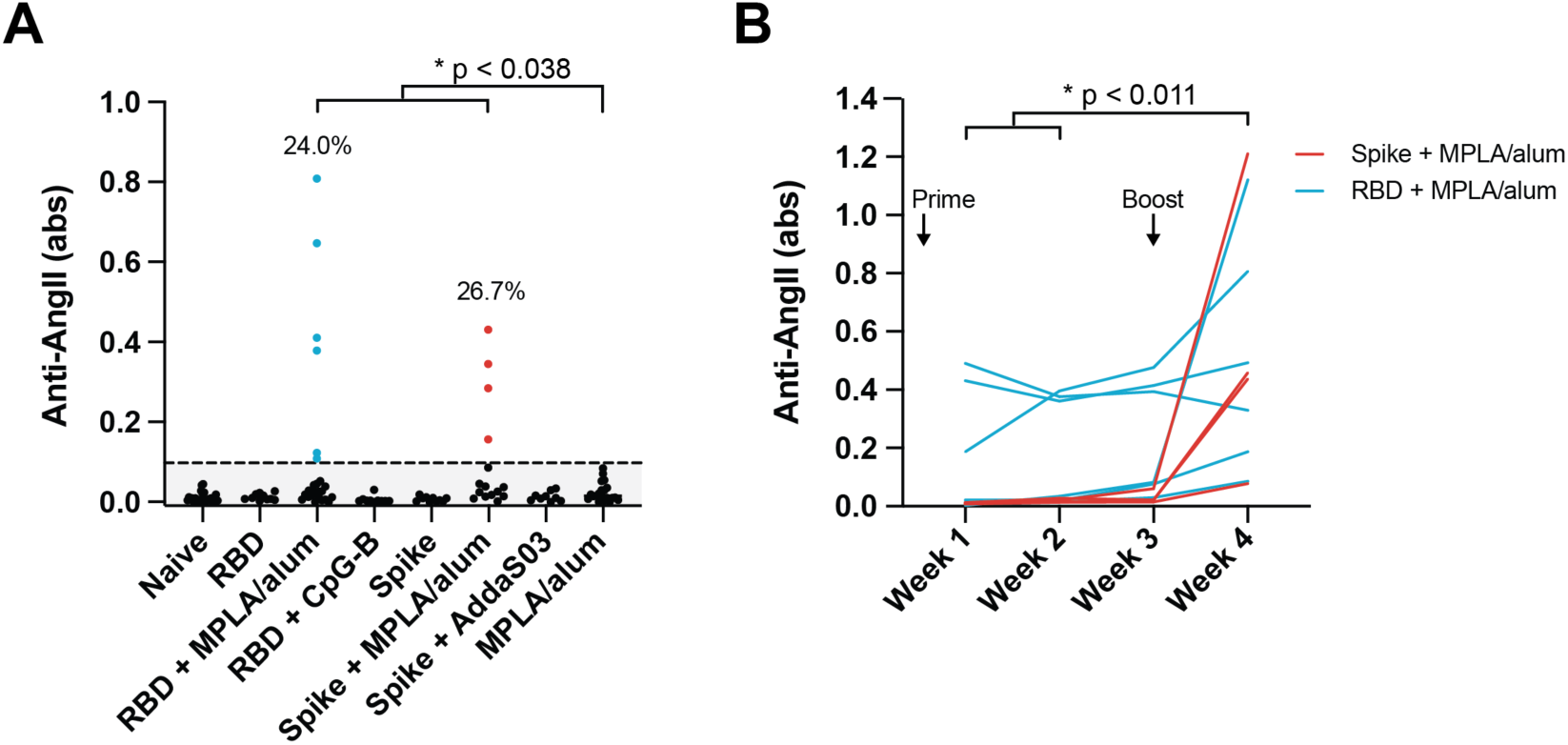
Development of anti-AngII autoantibodies in a subset of mice vaccinated with adjuvanted SARS-CoV-2 antigens. Mice were vaccinated at week 0 and 3 with Spike or RBD proteins adjuvanted with MPLA/alum, CpG-B, AddaS03 or with no adjuvant, or with MPLA/alum only (in absence of SARS-CoV-2 antigen). N=10-25 mice per group. **(A)** Anti-AngII levels in vaccinated mice plasma at week 4. Significant proportions of mice vaccinated with RBD + MPLA/alum or Spike + MPLA/alum raised autoantibodies against AngII (Z-test for proportions: RBD+MPLA/alum vs MPLA/alum: Z=2.35, Spike+MPLA/alum vs. MPLA/alum: Z=2.45, grey threshold = limit for anti-AngII positivity). **(B)** Levels of anti-AngII autoantibodies over time (at week 1, 2, 3 and 4) in the mice that were found positive for anti-AngII at week 4 (Friedman test with Dunn’s post-test).

We further analyzed the time course of the immune responses in mice, measuring anti-AngII levels at weeks 1, 2, 3 and 4 for the mice vaccinated with RBD + MPLA/alum or Spike + MPLA/Alum that were positive for anti-AngII. This analysis highlighted an increase in anti-AngII levels over time, specifically between weeks 1-2 and week 4 post-vaccination **(Fig. 2B)**. Of note, the increase in anti-AngII IgG titers was more substantial after the vaccination boost at week 3. These results demonstrate that anti-AngII autoantibodies can be induced in mice by vaccination against Spike or RBD, in the absence of SARS-CoV-2 infection, at least with MPLA/Alum-containing adjuvants.

Having determined that anti-AngII antibodies could develop both in hospitalized COVID-19 patients and in mice after vaccination with MPLA/alum-adjuvanted SARS-CoV-2 antigens, we further examined hospitalized COVID-19 patients for potential correlative effects of anti-AngII antibodies on blood pressure dysregulation, blood oxygenation, and disease severity. First, we observed that the levels of anti-AngII were significantly higher in patients with dysregulated blood pressure (BP), as defined by (i) episodes of hypotension that required administration of vasopressors, (ii) large daily fluctuations in blood pressure (daily mean arterial pressure range (ΔMAP) ≥ 70 mmHg), or (iii) at least 2 consecutive days of hypotension (MAP < 65 mmHg) in patients with pre-existing HTN **(Fig. 3A)**. Moreover, this correlation remained significant when each of these subgroups were analyzed separately, despite the low number of patients available **(Fig. 3B)**. Of note, these 3 subgroups of patients with dysregulated PB are not exclusive, with 52% of patients belonging to multiple categories. Consistent with these observations, a majority of patients with blood pressure dysregulation were positive for anti-AngII (81% vs. 53% in patients with normal blood pressure, **Fig. 3C**), among which 45% had high levels of anti-AngII. And finally, patients needing vasopressors were the most likely to have positive or high levels of anti-AngII, although among each of the groups with dysregulated blood pressure, >80% were anti-AngII(+) **(Fig. 3D)**.

**Figure 3.**
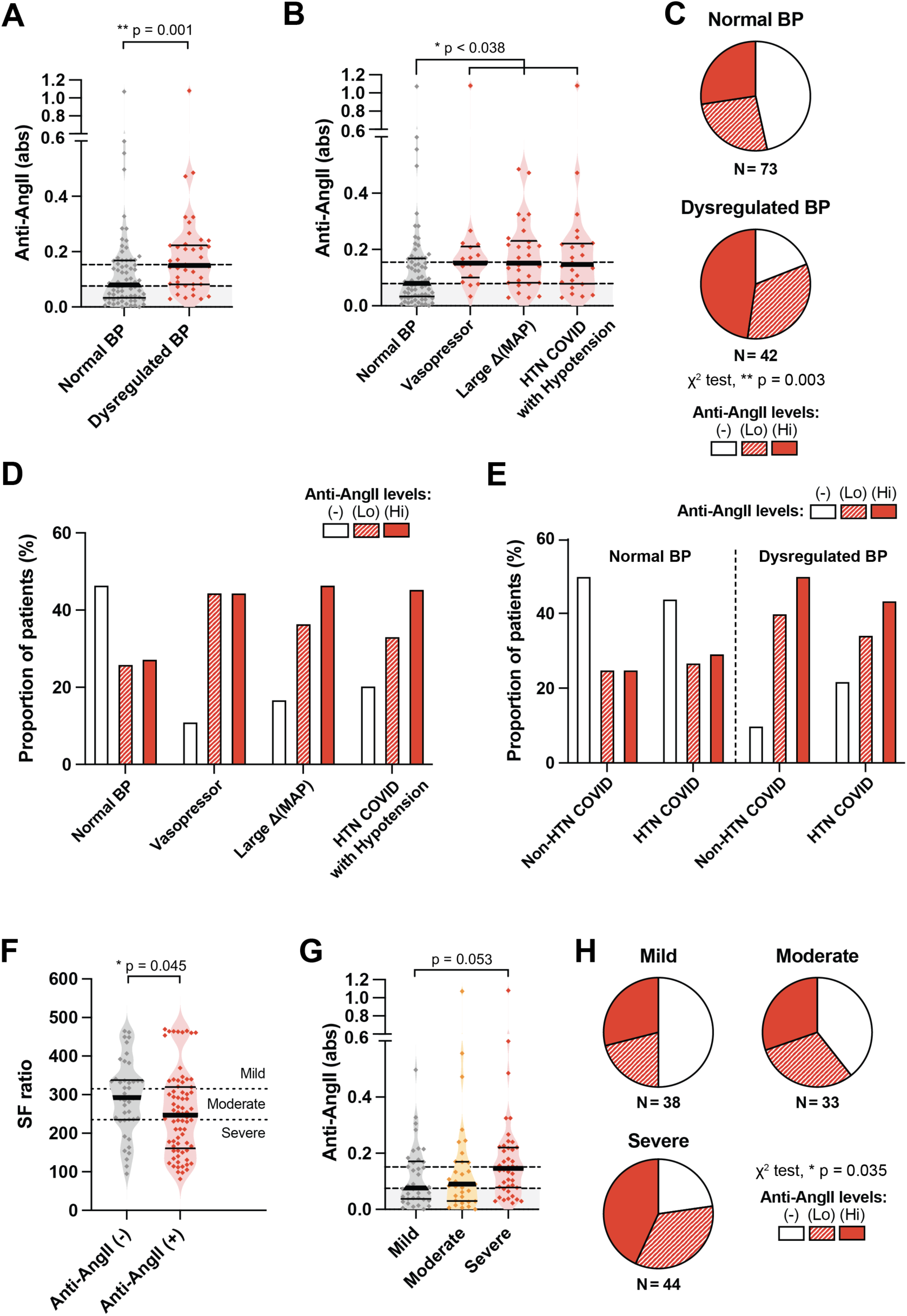
Anti-AngII autoantibodies correlates with dysregulated blood pressure (BP) and reduced pulse oxymetric saturation SpO_2_/FiO_2_ (SF ratio) in hospitalized COVID patients (N=115). Grey threshold indicates positivity of anti-AngII. **(A)** Anti-AngII levels in the plasma of COVID patients with normal or dysregulated blood pressure (Mann-Whitney test). **(B)** Levels of anti-AngII antibodies in COVID patients with normal BP as compared to patients with dysregulated BP, categorized as being under vasopressive drugs, exhibiting a large mean arterial pressure variability (large ΔMAP) or having experienced severe acute hypotensive episodes despite their known pre-existing HTN condition (these categories are non-exclusive; Kruskal-Wallis test with Dunn’s post-test comparison to normal BP). **(C)** Proportion of patients with normal or dyregulated BP that have negative (-), low (Lo) or high (Hi) levels of anti-AngII antibodies (χ^2^ test). **(D)** Proportions of patients with negative, low or high levels of anti-AngII in the COVID patients categories described in panel B. **(E)** Proportions of patients with negative, low or high levels of anti-AngII in COVID patients with pre-existing HTN and/or dysregulated BP. **(F)** SF ratio of COVID patients that are negative (-) or positive (+) for anti-AngII autoantibodies (Mann-Whitney test). SF ratio values are used to define disease severity as being mild, moderate or severe. **(G)** Levels of anti-AngII antibodies in patients suffering from mild, moderate or severe form of COVID, as defined based on SF ratio values (Kruskal-Wallis test with Dunn’s post-test). **(H)** Proportion of patients with negative (-), low (Lo) or high (Hi) levels of anti-AngII antibodies in function to the severity of the disease (χ^2^ test).

Since AngII is a critical regulator of blood pressure, we further hypothesized that COVID-19 patients with pre-existing HTN might be more sensitive to the effects of anti-AngII antibodies, if they were sufficiently abundant to alter the angiotensin signaling cascade. Therefore, we separated the patients with normal or dysregulated BP into subsets of patients with or without pre-existing HTN. Nevertheless, we found no striking difference between the non-HTN vs. HTN subsets that could support enhanced effects of anti-AngII in COVID patients with pre-existing HTN **(Fig. 3E)**.

Next, we questioned whether the presence or levels of anti-AngII antibodies correlated with reduced blood oxygenation and disease severity, measured via the daily mean oximetric saturation (SF ratio), which is the ratio of peripheral oxygen saturation (SpO2) to the fraction of inspired oxygen (FiO2). For each patient, we selected the lowest daily mean SF ratio value obtained within a two-day window of the measurement of anti-AngII. The lowest daily mean SF ratio was compared between the COVID-19 patients who developed auto-antibodies against AngII (73 patients) and those who did not (42 patients). We found that the SF ratio was strongly reduced in anti-AngII(+) patients (**Fig. 3F**), demonstrating a correlation between anti-AngII antibodies and reduced blood oxygenation in these patients.

Since the clinical severity of COVID directly depends on the patient’s blood oxygenation, patients with SF ratio ≥ 314 are considered as having a disease of low severity, while patients with SF ratio < 235 suffer from a highly severe form of COVID. Based on this, we then compared anti-AngII levels with disease severity and found that patients with severe vs. mild disease had increased anti-AngII antibodies (p=0.053, **Fig. 3G**). Similarly, we found a significant increase in the proportion of anti-AngII(+) patients along with disease severity (p=0.035 by χ^2^ test, **Fig. 3H**). Indeed, 77% of patients with severe disease were anti-AngII(+), including 41% with high levels, as compared to 50% and 29%, respectively, in patients with mild disease.

Together, these results demonstrate a correlation between the presence and levels of anti-AngII in COVID-19 patients and dysregulated blood pressure, lower blood oxygenation, and increased disease severity. While causal relationships cannot be drawn from these correlations, the fact that AngII is a key regulator of blood pressure in humans suggests that antibodies directed against anti-AngII could alter AngII signaling pathways, including its binding to and signaling via the angiotensin receptors AT1 and AT2. as well as on its enzymatic conversion by ACE2. Further experiments are needed to establish the pathophysiological consequences of anti-AngII auto-antibodies in COVID-19 patients.

Because anti-AngII antibodies developed in response to SARS-CoV-2 infection in hospitalized patients or vaccination with some adjuvants in mice, we hypothesized that anti-AngII antibodies may result from molecular structural mimicry between the AngII peptide and certain epitopes present in Spike or RBD. In that case, anti-AngII and anti-RBD could potentially cross-bind to both antigens. To test this hypothesis, we assessed the binding of two murine IgG monoclonal anti-AngII antibodies (clone E7 and clone B938M) to recombinant SARS-CoV-2 Spike or RBD. We found that both monoclonal anti-AngII antibodies bound to Spike and RBD, although with a lower affinity to RBD than to Spike **(Fig. 4A)**. We further demonstrated that the monoclonal anti-AngII antibodies may interfere with AngII binding to its cognate receptor AT1, suggesting that AngII-AT1 signaling could be modulated by the presence of anti-AngII **(Fig. 4B)**.

**Figure 4.**
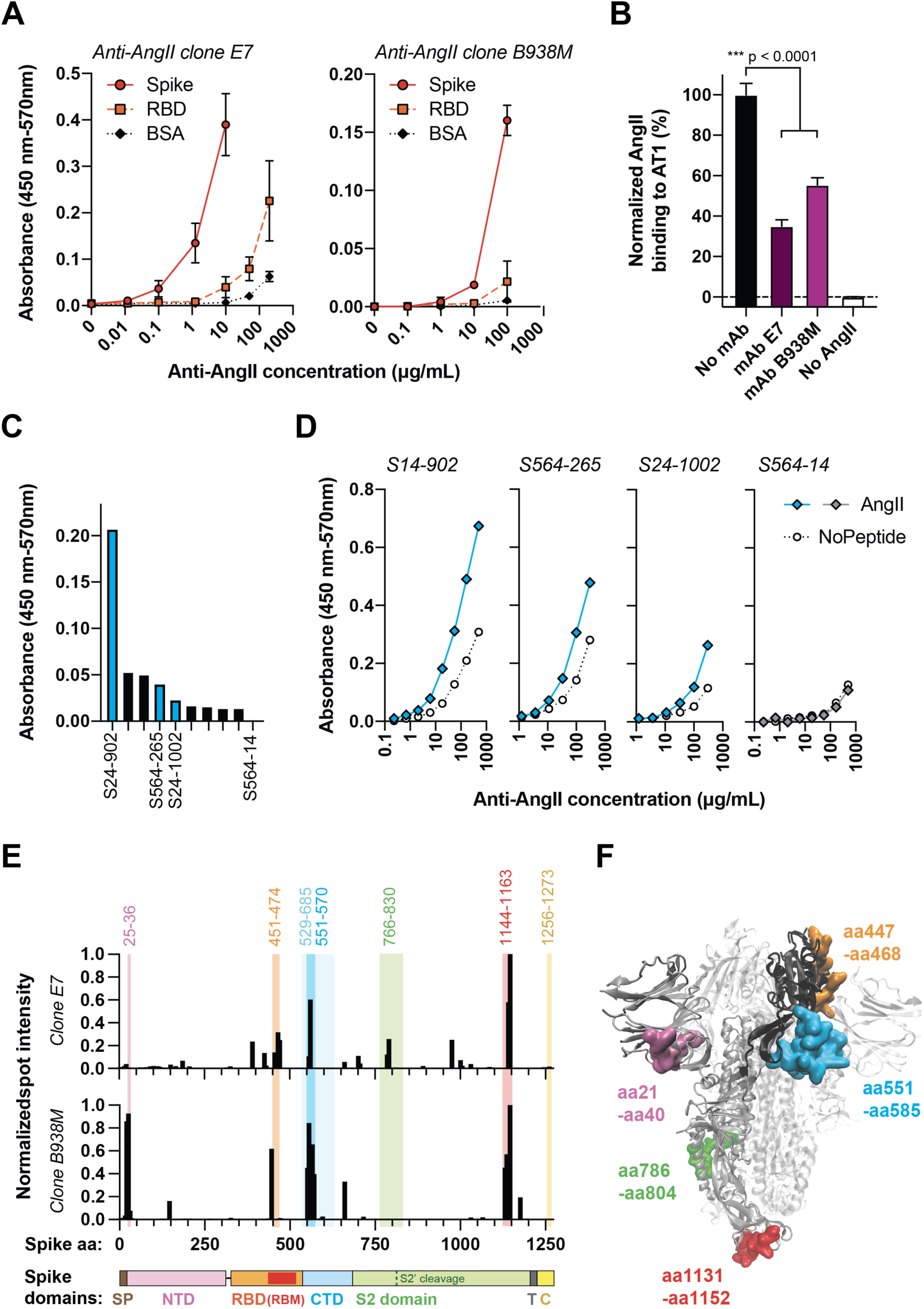
Cross-binding of anti-AngII antibodies to the SARS-CoV-2 Spike and RBD antigens. **(A)** Binding of two commercial monoclonal anti-AngII (clone E7 and clone B938M) to recombinant Spike and RBD at various concentrations by ELISA. **(B)** Inhibition of AngII binding to its cognate receptor AT1 in presence of the anti-AngII (clone E7 and B938M) on CHO-AT1 cells, measured by flow cytometry (ANOVA test with Tukey’s post-test). **(C)** Binding of a library of monoclonal anti-RBD antibodies against AngII by ELISA (9 out of 36 tested showed signals of low binding). **(D)** Binding of 3 selected monoclonal anti-RBD (S24-902, S564-265 and S24-1002) to AngII at various concentrations as compared to a non-AngII-binding anti-RBD (S564-14). **(E)** Binding of monoclonal anti-AngII (clone E7 or B938M) to Spike linear epitopes using a peptide array. Highlighted colored regions represents the most immunogenic and recurrent B cell epitopes of Spike found in COVID patients (Shrock *et al*. (*12*), Li *et al*. (*13*)) and the associated numbers indicate the amino-acid positions that delineate these regions. The x-axis represents the full-length of Spike (SP = signal peptide; NTD = N-terminal domain; RBD = receptor-binding domain; RBM = receptor-binding motif; CTD = C-terminal domain; NTD, RBD and CTD together constitutes the S1 domain of Spike; T = transmembrane domain; C = cytoplasmic tail). **(F)** Visualization of the main domains targeted by the monoclonal anti-AngII antibodies on the 3D structure of Spike (PDB=6VXX, (*14*)). Domains are highlighted in colors on one monomer of the trimeric Spike structure (RBD is in black; aa# = amino acid position of Spike).

Conversely, we evaluated the ability of monoclonal anti-RBD antibodies, in a library of those isolated from SARS-CoV-2 infected patients, to bind to AngII. Among the 36 different monoclonal anti-RBD antibodies assessed, 9 showed some low to very-low binding to AngII, with one being superior to the others, namely clone S24-902 **(Fig. 4C)**. We further confirmed the significant although low binding of 3 of these anti-RBD to AngII, respectively S24-902, S564-265 and S24-1002, by characterizing their affinity at various concentrations **(Fig. 4D)**. In contrast, clone S564-14 did not show any specific binding to AngII and was used as a negative control. It is not surprising that the cross-reactivity of anti-RBD to AngII is low, considering that monoclonal anti-AngII also binds weakly to RBD **(Fig. 4A)** and that AngII is a short 8-amino acid peptide.

We then sought to identify the epitopes of Spike or RBD that lead to cross-reactivity to AngII. To do so, we tested the binding of the two monoclonal anti-AngII antibodies (clones E7 and B938M) to linear epitopes of Spike, using a peptide array that consists of a library of 15-mer peptides, shifted by 5 amino acids, covering the full-length of Spike. As a control for nonspecific binding, we used the secondary antibody only, in the absence of anti-AngII antibody. Both anti-AngII E7 and B938M clones bound to several linear epitopes on Spike, with a main target at Spike residues aa1146-aa1160 located near the C-terminus, outside of RBD (**Fig. 4E, Fig. S3A-C**, spot J14 on the peptide array), and about 15 secondary targets of lower affinity. Five of the secondary peptides targeted by the clone E7 belong to the RBD of Spike, including 3 more particularly to the receptor-binding motif (RBM) of the RBD **(Fig. 4E, Fig. S3A)**. In contrast, only 1 peptide targeted by the B938M clone belongs to the RBD region, in the RBM **(Fig. 4E, Fig. S3B)**. Of note, although both Spike and AngII share the same receptor, ACE-2, their binding on ACE-2 occurs at 2 distinct locations **(Fig. S4)** (*10, 11*). Importantly, most regions targeted by clones E7 and/or B938M have been characterized as being the main B cells epitopes in COVID patients (colored regions in **Fig. 4E, F, Fig. S3D**) (*12, 13*). These regions, which are likely to contain the epitopes generating anti-AngII autoantibodies, are (1) aa21-aa40, (2) aa447-aa468, (3) aa551-aa585, (4) aa786-aa804, (5) aa1131-1160 of Spike.

We next compared the Spike epitopes targeted by polyclonal responses raised against RBD/Spike in vaccinated mice or in COVID patients to the ones targeted by the monoclonal anti-AngII. Thus, we repeated the peptide array assays using anti-AngII(+) or anti-AngII(-) pooled sera from 5 different mice vaccinated with Spike + MPLA/alum or RBD + MPLA/alum, or from 5 different COVID-19 patients.

We observed that mice raised anti-Spike/RBD antibodies in all regions previously targeted by the monoclonal anti-AngII upon vaccination with Spike+MPLA/alum **(Fig. S5A)**, while mostly in the region aa447-aa468 upon vaccination by RBD+MPLA/alum **(Fig. S5B)**. The profiles of anti-AngII(+) and anti-AngII(-) mice were very similar, except for a small increase in binding of anti-Spike to the region aa447-aa468 and anti-RBD to aa551-585, in anti-AngII(+) mice. As to the COVID-19 patients, they raised anti-Spike antibodies against 3 out of the 5 anti-AngII-targeted regions, namely aa551-aa585, aa786-aa804 and aa1131-1160, with a slightly higher binding observed to aa786-aa804 in anti-AngII(+) plasma **(Fig. S5C)**. Surprisingly, the region aa1131-1160 was better targeted in anti-AngII(-) plasma; this highlights that not every antibody that bind to anti-AngII-targeting regions would necessarily cross-react with AngII.

Taken together, using monoclonal anti-AngII and anti-RBD antibodies, we demonstrate that antibody cross-binding between AngII and Spike/RBD can occur, even if weakly, which may suggest some structural homology between AngII and certain epitopes of Spike/RBD.

SARS-CoV-2 infection has been shown to induce broad immune dysregulation (*15, 16*), including generation of wide-ranging autoantibody responses (*17*). For example, generation of autoantibodies against phospholipids has been shown to contribute to coagulation disorders (*18, 19*), and against Type I interferons to reduced immune response to infection (*20*). Here, we show that autoantibodies can be generated against AngII, a key regulator of vascular tension. We further show that generation of anti-AngII autoantibodies correlated with disease severity as reflected in dysregulation of vascular tension and lower blood oxygenation. This, along with our *in vitro* signaling data, suggests that the anti-AngII autoantibodies, even if low affinity, may be able to interfere with signaling between AngII and its receptors AT1 and ACE-2. Interestingly, some COVID-19 patients had also been shown to develop autoantibodies against AT1 and ACE-2, which similarly correlated with enhanced pro-inflammatory responses and increased disease severity (*21, 22*). Such autoantibodies against AT1 and ACE-2 have been also observed in patients suffering from other vascular pathologies, particularly in malignant hypertension (*8*) or constrictive vasculopathy (*9*). Therefore, systematic quantification of autoantibodies against key molecules of the renin-angiotensin pathway (i.e. AngII, AT1, and ACE-2), in diseases characterized by vascular dysregulation, including COVID-19, might reveal a common underlying autoimmune etiology. Lastly, we highlighted the immune epitopes of Spike that could share structural homology with AngII, providing molecular insights in the immune mechanisms that could lead to the generation of anti-AngII autoantibodies upon infection by SARS-CoV-2.

## Data Availability

All data produced in the present work are contained in the manuscript.

## Acknowledgements

The authors would like to thank all members from the laboratories of Prof. Jeffrey A. Hubbell and Prof. Melody A. Swartz that participated on COVID research, Yuanyuan Zha, Hongyuan Jiao, Glee Guilan Li and Shuhan Yu for biobank COVID sample processing, Kelly M. Blaine for collecting samples from hypertensive and control donors and Prof. Prabha Siddarth (University of California Los Angeles, CA USA) for advice on data analysis.

## Funding

This study was funded by the University of Chicago, through the Chicago Immunoengineering Innovation Center, the Chicago Biomedical Consortium and the University of Chicago’s “Big Ideas Generator” for COVID research. S.R.J. was supported by the T32 CA009566.

## Authors contributions

P.S.B, J.A.H., M.A.S designed the study, analyzed the data and wrote the manuscript. P.S.B. has performed the experiments. S.J.R., J.Y., A.R.P., J.T., T.F.G. and A.I.S collected patient samples and clinical data. S.J.R. processed and analyzed clinical data. H.L.D., C.T.S., S.C. and P.C.W. produced, purified, characterized and provided the monoclonal anti-RBD antibodies. All authors have reviewed the manuscript.

## Competing interests

P.S.B., J.A.H. and M.A.S. are named as inventors on US Provisional Patent 63/123,199, filed December 9, 2020, relating to measurement of anti-AngII in immune response. The University of Chicago has filed a patent application relating to anti-SARS-CoV-2 antibodies generated by P.C.W., H.L.D., and C.T.S. as co-inventors.

## Supplementary Materials

Materials and Methods

Fig. S1-S5

## Notes

### Author Declarations

The IRB of the University of Chicago, Department of Medicine gave ethical approval for this work.

