## Supplementary material for "SARS-CoV-2 infection induces cross-reactive autoantibodies against angiotensin II": Materials and Methods / Supplementary Figures

**This PDF file includes:**

Materials and Methods
Figs. S1 to S5

### Materials and Methods

Human plasma biobank and clinical data collection

All samples collection and data recording on COVID-19 patients were approved by the IRB, under an informed consent, from a cohort at the University of Chicago. Serum samples were also collected from both non-hypertensive and hypertensive organ donors provided by the Gift of Hope Organ Donor Network (Itasca, IL). Patients of the COVID cohort were hospitalized patients, in contrast to non-SARS-CoV-2 infected healthy and hypertensive individuals, which were non-hospitalized donors. In addition, many of the COVID-19 patients had pre-existing health conditions, including hypertension. Blood was analyzed for the presence and amounts of specific anti-RBD IgG and anti-AngII IgG antibodies by enzyme-linked immunosorbent assay (ELISA).

Titration of anti-RBD antibodies

All procedures utilizing human samples were performed under Biosafety level 2 conditions. For the titration of anti-RBD antibodies, ELISA plates were coated with purified recombinant RBD (100 nM) in PBS for 12-16 hours at 4°C. Plates were then blocked with bovine serum albumin (BSA, 2%) in PBS for 2 hours at room temperature. Plates were then extensively washed with PBS-Tween 0.05% (PBST). Human plasmas were diluted at 1:100 in PBST + BSA (0.5%), and then serially diluted by 10. Samples were added to the plates and incubated for 2 hours at room temperature. Plates were washed again and incubated with a horseradish peroxidase (HRP)-conjugated anti-human IgG for 1 hour at room temperature in the dark. Plates were washed, revealed using 3,3',5,5'-Tetramethylbenzidine (TMB) for 30 min in the dark, and the reaction was stopped using 1M $H_2SO_4$. Absorbances at 450 nm were read using an Epoch plate reader (BioTek), and corrected with the absorbance at 570 nm. Titers represent the highest dilution at which antibodies were detected, in log10 (e.g. an IgG titer of 4 indicates a detectable level of anti-RBD IgG in a plasma sample diluted at 1:10000). A value of 0 was given to samples that had no detectable IgG at a dilution of 1:100. Anti-RBD titers are reported in $\log_{10}$. Patients were considered positive for anti-RBD when their titers was ≥ 4, since some healthy donors samples taken prior to the COVID-19 pandemic had titers of 3. Anti-RBD titers were considered low at 4-5 and high at 5-8.

Detection of anti-AngII antibodies

For the detection of anti-AngII antibodies, streptavidin-coated plates were purchased from Pierce (ThermoFisher Scientific) and biotin-LC-AngII was purchased from AnaSpec. Streptavidin-coated plates were re-hydrated using PBST, and biotin-LC-AngII (10 μg/mL) was added to the plate for 2 hours at room temperature. Plates were washed with PBST, and plasma samples diluted at 1:100 were added to the plates for 2 hours at room temperature. Plates were washed again and incubated with an HRP-conjugated anti-human IgG for 1 hour at room temperature in the dark. Plates were washed and developed as previously described, except that the signals were stopped after about 10-15 min (when the absorbance of the reference well reached ~ 1 AU). Absorbances at 450 nm were corrected with the absorbance at 570 nm. Detection of antibodies in absence of biotin-LC-AngII was also performed and subtracted to correct for AngII-unspecific antibody binding. Threshold to define positive signals is 3 standard deviations above the mean of the healthy donors cohort (after removal of the outliers according to the IQR rule) which corresponds to an absorbance of 0.077. Threshold to define high levels of anti-AngII is twice the threshold for positivity, which is an absorbance of 0.154.

Data analysis of anti-AngII and anti-RBD in COVID patients

Many patients had several plasma samples available at different days post-symptoms onset (DPSO). Anti-AngII and anti-RBD levels were measured for all available timepoints. As a rule, the highest value of anti-AngII for each patient was selected for data analysis, along with its associated anti-RBD titers at the same timepoint. The analysis of the anti-AngII/anti-RBD levels changes between early and late timepoints was performed on the 25 patients that were positive for anti-AngII and had samples availables for both time ranges in 1-10 and 11-21 DPSO. Again, the highest anti-AngII values in 1-10 and 11-21 DPSO were selected when multiple timepoints were available, and the anti-RBD corresponded to corresponding timepoints of highest anti-AngII. For the analysis at > 45 DPSO, the highest anti-AngII value detected after 45 DPSO was similarly selected when more than one sample was available for a patient.

The categories for the body mass index (BMI) analysis were chosen according to the Center for disease control and prevention (CDC) definitions: underweight = BMI < 18.5; normal = BMI between 18.5 and < 25; overweight = BMI between 25 and < 30; obese = BMI between 30 and < 40; and severly obese = BMI ≥ 40.

Data analysis of anti-AngII correlations to blood pressure dysregulation, blood oxygenation and disease severity

Patients that were considered having a dysregulated blood pressure were: (1) the ones that received vasopressive drugs at any time during their hospitalization, (2) the ones that had large daily fluctuation of mean arterial pressure range ΔMAP ≥ 70 mmHg or (3) the ones that had pre-existing hypertension (HTN) and experienced at least 2 consecutive days of acute hypotension (MAP < 65 mmHg). For the patients in (2) and (3), the dysregulation of blood pressure was assessed in the ± 3 days around the timepoint of the highest anti-AngII value of the patient. Similarly, the SF ratio selected for each patient was the lowest SF ratio value detected in the ± 3 days around the timepoint of the patient's highest anti-AngII value. Disease severity was defined according to the SF ratio value selected for each patient as such: mild disease = SF ratio ≥ 315; moderate disease = SF ratio between 235 and < 315; severe disease = SF ratio < 235.

Protein productions

The ectodomain of Spike (BEI Resources: NR-52310) or the subdomain RBD (BEI Resources: NR-52309) were cloned into pCAGGS vector for mammalian expression in human embryonic kidney (HEK) 293F suspension cells, as his-tagged recombinant proteins. The proteins were expressed for 4-7 days in FreeStyle medium at 37°C, 5% $CO_2$, under constant agitation. Cell supernatants were then collected and filtered at 0.22 µm. The proteins were purified by immobilized nickelaffinity chromatography (HisTrap HP column, GE Healthcare) using fast protein liquid chromatography (FPLC; Äkta Pure system, GE Healthcare). Some protein batches were additionally purified by size exclusion chromatography (Superdex 200 pg column, GE Healthcare), and/or added with dithiothreitol (DTT) 10 mM to reduce protein dimerization. Proteins were then extensively dialyzed against phosphate-buffered saline (PBS; pH 7.4), sterile-filtered and stored at -80°C. All proteins were tested for endotoxin level < 2 EU/mg.

<u>Vaccines formulation</u>

Vaccines were formulated in PBS, by mixing 10 µg SARS-CoV-2 antigens, i.e. RBD or Spike, with various adjuvants. MPLA/alum was prepared by mixing 5 µg of MPLA (InvivoGen) with 50 µg of Alum (InvivoGen). AddaS03 was prepared by mixing 25 µL of AddaS03™ (Invivogen) with 25 µL of PBS containing the SARS-CoV-2 antigens. Mouse specific stimulatory CpG class B (ODN1826; InvivoGen) was injected at a dose of 20 µg. Non-adjuvanted groups were composed of 10 µg of antigen only. Mice vaccinated with MPLA/alum without antigens were injected with 5 µg of MPLA and 50 µg of Alum.

<u>SARS-CoV-2 vaccination and analysis in mice</u>

All mouse experimentation was approved by the IACUC at the University of Chicago. Female 8-week old C57BL/6 mice received a prime vaccination using the vaccine formulations above-described, as well as a boost vaccination 3 weeks later. Vaccinations were administered by intradermal injections in the animal hocks in the two forelimbs (25 µL/hock). Mice were bled weekly via the submandibular vein after vaccination for plasma collection and analysis. Anti-AngII IgG levels were determined as described above, except using an anti-Ms total IgG as a detection antibody (1:8000; Southern Biotech). Threshold to define positive signals is 3 standard deviations above the mean of the group injected with MPLA/alum without SARS-CoV-2 antigen, which is an absorbance of 0.095.

Binding of monoclonal anti-AngII antibodies to RBD and Spike

A mouse monoclonal IgG2a anti-AngII/III clone E7 (MA1-82996) was purchased from ThermoFisher Scientific and a mouse monoclonal IgG1 anti AngI/II/III (GTX44411) was purchased from GeneTex. ELISA plates were coated with RBD or Spike (100 nM) in PBS for 12-16 hours at 4°C. The plate was then blocked using BSA 2% for 2 hours at room temperature. The anti-AngII antibody was diluted in PBST-BSA (0.5%) at the specified concentrations and added to the plate for 2 hours at room temperature. Binding of the anti-AngII to RBD, Spike or BSA was detected using an anti-mouse IgG (Southern Biotech) for 1 hour at room temperature in the dark. The plate was then revealed and the absorbance determined as described above.

Inhibition of AngII binding to AT1 by anti-AngII monoclonal antibodies

Binding of AngII to AT1 receptors was assessed using fluorescently labelled FAM-AngII, purchased from AnaSpec, and Chinese Hamster Ovarian (CHO) cells expressing recombinant human AT1 (CHO-AT1 cells), purchased from Perkin-Elmer. FAM-AngII (100 nM) was pre-incubated with 0.3 mg/mL of the anti-AngII monoclonal antibodies (clone E7 or B938M) in PBS for 30 min at 37°C or with no antibody as a negative control. Then, CHO-AT1 cells were stained with these mixtures in the dark for 20 min on ice. Cells were washed 3 times in PBS + 2% Fetal Bovine Serum, after what the binding of FAM-AngII to AT1 on the cell surface was detected using a flow cytometer (BD LSRFortessa, BD Biosciences). The mean fluorescence intensity (MFI) of the FAM-AngII was computed using FlowJo software.

Binding of monoclonal anti-RBD to AngII

A library of monoclonal anti-RBD was isolated and sequenced from COVID-19 patients, produced as recombinant proteins using HEK-293 cells and purified using Protein-G affinity-based purification. The binding of the monoclonal anti-RBD to AngII was assessed as described above, by coating streptavidin-coated plate with 10 µg/mL biotin-LC-AngII, washing with PBST, incubating with anti-RBD clones at

20 µg/mL or indicated concentrations, washing again, and detecting with an anti-human IgG. Negative controls were done in absence of biotin-LC-AngII.

Peptide array of SARS-CoV-2 Spike

SARS-CoV-2 Full Spike CelluSpots peptide array assays were purchased from Intavis Bioanalytical Instruments. The assay contains 254 peptides of 15 amino acids (aa) length covering the full-length Spike sequence with a shift of 5 aa between peptides. The assay was performed as instructed by the manufacturer. Briefly, the membrane was blocked with casein blocking buffer (Sigma-Aldrich) for 4 hours at room temperature, and then incubated for 12 hours at 4°C with the anti-AngII clone E7 diluted at a concentration of 20 µg/mL in casein. The membrane was then washed extensively using PBST, and further incubated with an HRP-conjugated anti-mouse IgG (1:5000; Southern Biotech) for 2 hours at room temperature. The membrane was again extensively washed in PBST and revealed using the Clarity ECL Western substrate (BioRad). The membrane was imaged using a gel imager (BioRad) and spot intensity were analysed using the Protein Array Analyze for ImageJ (2010) made by Carpentier G. and available online at: http://rsb.info.nih.gov/ij/macros/toolsets/Protein Array Analyzer.txt. For peptide arrays using plasma from COVID-19 patients or vaccinated mice, the same procedure was followed, except that the pooled plasma from 5 individuals was diluted at 1:200 in casein and incubated for 4 h at room temperature instead of the incubation with anti-AngII antibodies. For patients samples, an anti-human IgG was used as a secondary antibody to reveal signals.

Statistics and softwares

Graphs and statistical analysis were performed using Prism 9 (GraphPad Software LLC). Median ± interquartile range are represented in violin plots, while mean ± SD are represented in other types of graphs. Non-parametric tests (Mann-Whitney for 2 groups comparison, Wilcoxon for paired data, Kruskall-Wallis for >2 groups comparisons) were used for data with non-normal distribution. Spearman tests were used for correlations. ANOVA were used for multiple groups comparison on normally distributed data. All tests were double-sided and the p-values were corrected for multiple comparisons. Statistics for $\chi^2$ proportion tests were performed online using https://www.socscistatistics.com calculators. $\chi^2$ tests were used to compare the % of anti-AngII positive patients between the different categories of interest. Threshold for statistical significance was $p < 0.05$.

Excel (Microsoft) was used to sort and analyze the data. Molecular structures visualization rendering were done using VMD (Visual Molecular Dynamics 1.9.1). Illustrator CS5 (Adobe) was used to make the figures.

### Supplementary Figures

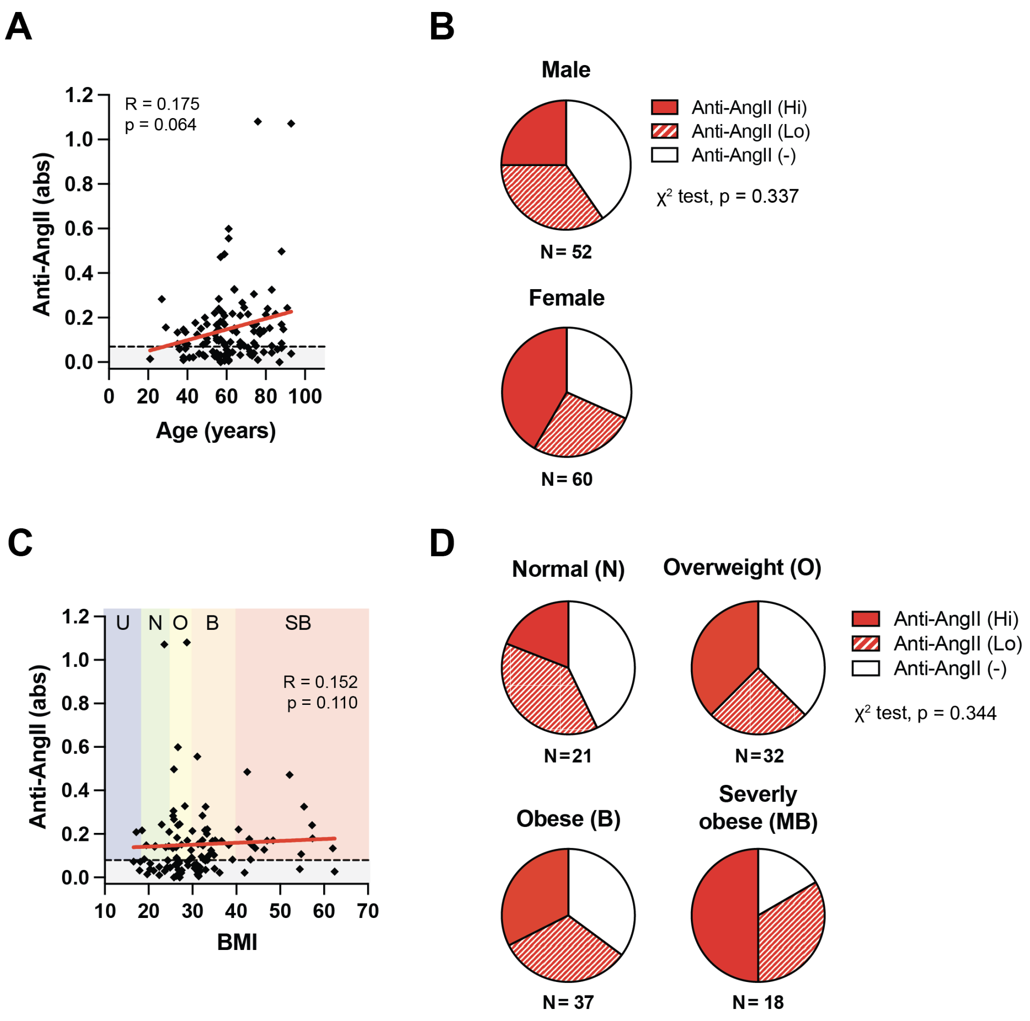


***Supplementary Figure 1.*** **Anti-AngII does not correlate with sex, age and body mass index (BMI) in hospitalized COVID patients (N=112).** Grey threshold = limit for anti-AngII positivity. **(A)** Correlation between the level of anti-AngII antibodies and the age of patients (Spearman correlation; linear regression in red). **(B)** Proportion of male or female patients with high (Hi), low (Lo) or negative (-) levels of anti-AngII ($\chi^2$ test). **(C)** Correlation between the level of anti-AngII antibodies and the BMI of patients (Spearman correlation; linear regression in red). **(D)** Correlation of the age of the patients with their level of anti-AngII (Spearman correlation; linear regression in black). U = underweight, N = normal weight, O = overweight, B = obese, SB = severely obese. **(E)** Proportion of patients of different weight categories that had no (-), low (Lo) of high (Hi) anti-AngII levels ($\chi^2$ test).

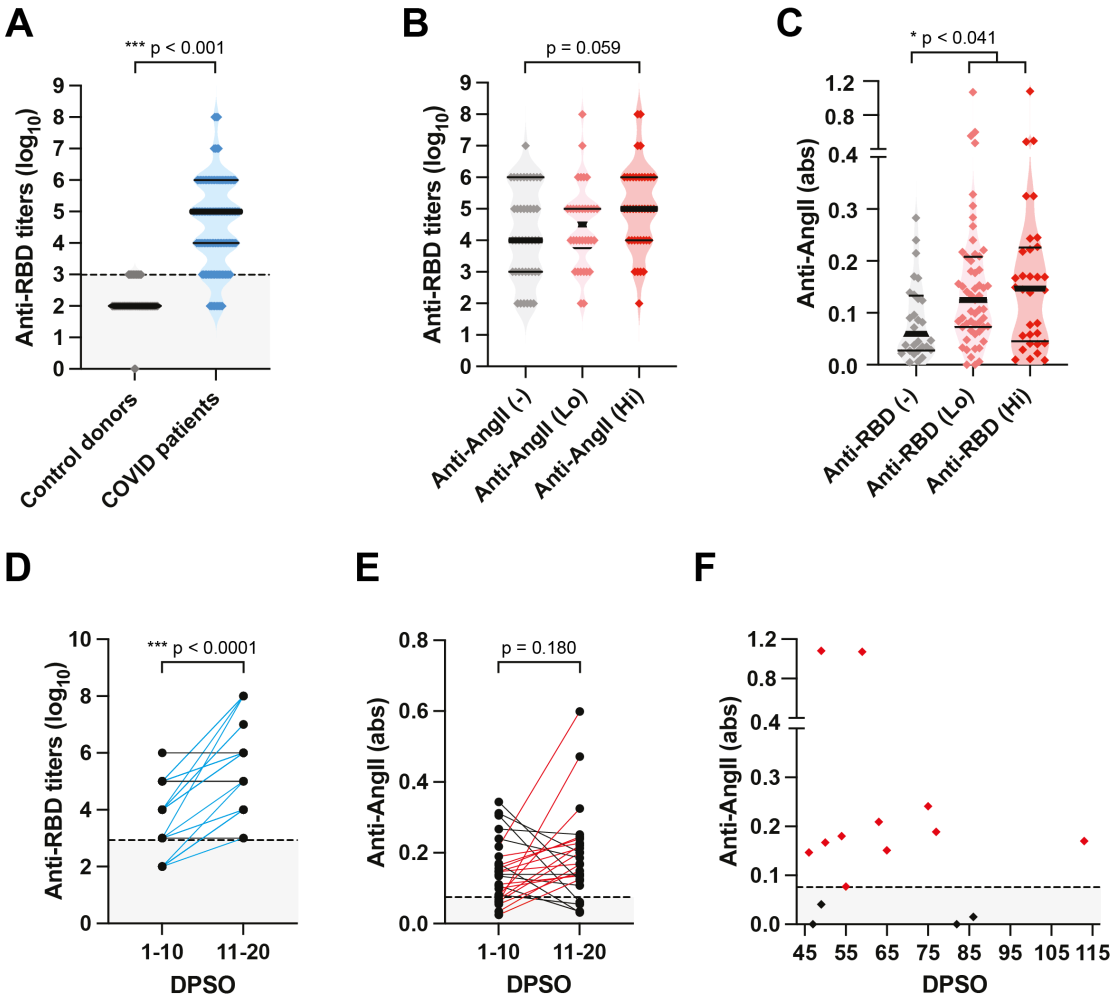


***Supplementary Figure 2.* Anti-AngII antibodies level does not strongly correlate with anti-RBD titers in COVID patients (N=115).** **(A)** Anti-RBD titers ($log_{10}$) in hospitalized COVID patients as compared to control donors (sampled before the COVID pandemy; Mann-Whitney test). **(B)** Anti-RBD titers in COVID patients with negative (-), low (Lo) or high (Hi) levels of anti-AngII (Kruskall-Wallis test with Dunn's post-test). **(C)** Level of anti-AngII in COVID patients with negative (-), low (Lo) or high (Hi) titers of anti-RBD (Kruskall-Wallis test with Dunn's post-test). **(D)** Increase in anti-RBD titers in COVID patients between 1-10 and 11-20 days post-symptoms onset (DPSO) (N=25; Wilcoxon matched pairs signed rank test). **(E)** Increase in anti-AngII levels in COVID patients between 1-10 and 11-20 DPSO (N=25; Wilcoxon matched pairs signed rank test). **(F)** Levels of anti-AngII at late time (>45 DPSO) after symptoms onset (N=15 patients).

**A**

**Spike peptide sequences detected by anti-AngII clone E7**

| | | | |
|---|---|---|---|
| **D7:** | CFTNVYADSFVIRGD | **F21:** | AENSVAYSNNSIAIP |
| **D14:** | PDDFTGCVIAWNSNN | **F22:** | AYSNNSIAIPTNFTI |
| **D20:** | FRKSNLKPFERDIST | **G14:** | KQIYKTPPIKDFGGF |
| **D22:** | RDISTEIYQAGSTPC | **G15:** | TPPIKDFGGFNFSQ |
| **D23:** | EIYQAGSTPCNGVEG | **I4:** | VLNDILSRLDKVEAE |
| **E16:** | NKKFLPFQQFGRDIA | **I9:** | LQSLQTYVTQQLIRA |
| **E17:** | PFQQFGRDIADTTDA | **J13:** | LQPELDSFKEELDKY |
| **F13:** | ECDIPIGAGICASYQ | **J14:** | **DSFKEELDKYFKNHT** |

**B**

**Spike peptide sequences detected by anti-AngII clone B938M**

| | | | |
|---|---|---|---|
| **A5:** | RTQLPPAYTNSFTRG | **E19:** | DTTDAVRDPQTLEIL |
| **A6:** | PAYTNSFTRGVYYPD | **F13:** | ECDIPIGAGICASYQ |
| **D18:** | GGNYNYLYRLFRKSN | **J11:** | GIVNNTVYDPLQPEL |
| **E15:** | VLTESNKKFLPFQQF | **J12:** | TVYDPLQPELDSFKE |
| **E16:** | NKKFLPFQQFGRDIA | **J14:** | **DSFKEELDKYFKNHT** |
| **E17:** | PFQQFGRDIADTTDA | **J20:** | VVNIQKEIDRLNEVA |
| **E18:** | GRDIADTTDAVRDPQ | | |

**C**

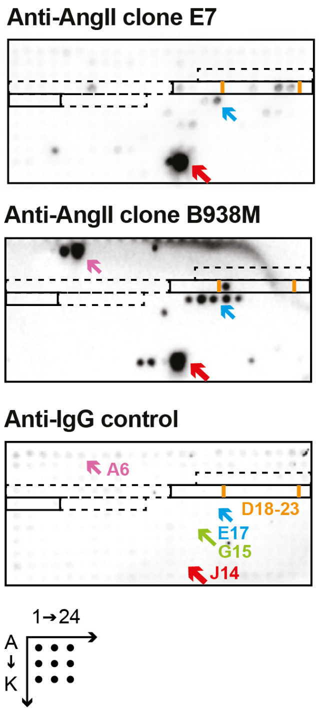


**D**

**Main anti-AngII targetted peptides:**

| | |
|---|---|
| aa21 - aa40: | **RTQLPPAYTNSFTRGVYYPD** |
| aa447 - aa468: | **GNYNYLYRLFRKSNLKPFERDI** |
| aa551 - aa585: | **VLTESNKKFLPFQQFGRDIADTTDAVRDPQTLEIL** |
| aa786 - aa804: | **KQIYKTPPIKDFGGFNFSQ** |
| aa1131 - aa1160: | **GIVNNTVYDPLQPELDSFKEELDKYFKNHT** |

***Supplementary Figure 3.*** **Epitopes targeted by the monoclonal anti-AngII clone E7 and B938M.** Monoclonal anti-AngII were added onto a peptide array containing the full-length of Spike with 15-mers peptide with a 5 amino-acids overlap. **(A)** Sequences of the Spike linear epitopes targeted by the anti-AngII clone E7. **(B)** Sequences of the Spike linear epitopes targeted by the anti-AngII clone B938M. **(C)** Representative images of the positive spots on the Spike peptide arrays probed with the anti-AngII clone E7 or B938M, or the anti-IgG secondary control only. The spot J14 is the primary target of both anti-AngII monoclonal antibodies. **(D)** Sequences of the main targeted domains by the anti-AngII monoclonal antibodies that are highlighted in the 3D structure of Spike in Fig. 4F, with the corresponding amino-acid positions (aa#). All domains are targeted by both anti-AngII clones except the aa21-aa40 domain, which is a strong target of the clone B938M only, and aa786-804 which is targeted by clone E7 only.

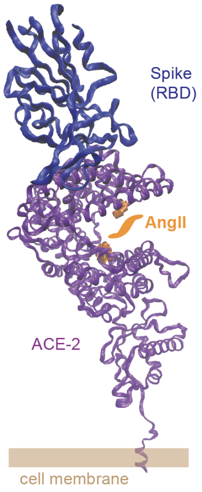


***Supplementary Figure 4.*** **Binding sites of SARS-CoV-2 RBD and AngII on the human Angiotensin Converting Enzyme (ACE)-2.** The binding of RBD on ACE-2 seems to occur at a different location than the catalytic site of AngII (PDB: 6M17 *(10)* by and AngII binding residues were determined from Guy *et al.* *(11)*).

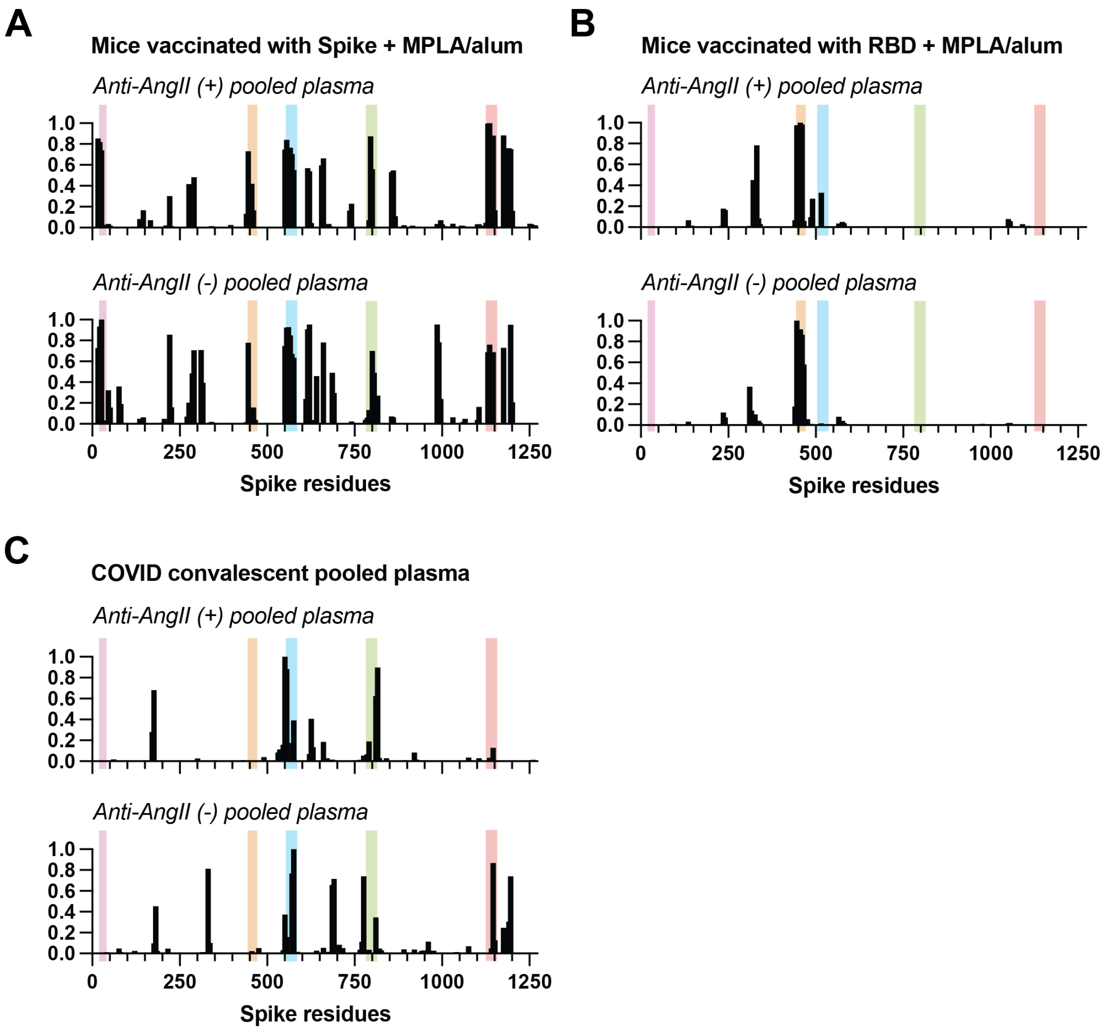


***Supplementary Figure 5.*** **The Spike linear epitopes targeted by anti-AngII monoclonal antibodies are the same as the ones targeted by the plasma IgG of mice vaccinated with recombinant Spike or RBD proteins or of COVID convalescent patients.** Highlighted colored regions are the main domains targeted by at least one anti-AngII monoclonal antibody. X-axes represent the position of linear epitopes covering the full-length of Spike. **(A, B)** Linear epitopes of Spike targeted by IgG antibodies in plasma of mice positive (+) or negative (-) for anti-AngII, upon immunization with recombinant Spike (A) or RBD (B) adjuvanted with MPLA/alum (plasma pooled from N=5 mice). **(C)** Linear epitopes of Spike targeted by IgG antibodies in plasma of COVID patients positive (+) or negative (-) for anti-AngII (plasma pooled from N=5 patients).
